# Dendritic cells and vascular inflammation in patients with coronary artery disease with and without chronic kidney disease

**DOI:** 10.64898/2026.10.01.26364549

**Authors:** Johannes Ruhe, Sonja Böttger, Tzvetanka Bondeva, Ilonka Rohm, Gloria Färber, Torsten Doenst, Daniel Kretzschmar, Gunter Wolf, Martin Busch

## Abstract

**Objective:** Dendritic cells (DCs) are priming local immune responses, and their tissue-associated activity increases in inflammatory conditions. In patients with coronary artery disease (CAD), circulating dendritic cell precursors (DCPs) are reduced. This study investigates circulating DCPs and tissue densities of migrated DCs in CAD patients undergoing bypass surgery, focussing on patients with chronic kidney disease (CKD) as a risk factor for CAD and a condition of chronic vascular inflammation.

**Approach and Results:** Circulating DCPs were measured in the peripheral blood (before surgery) using fluorescence-activated cell sorting (FACS) in n=59 patients and results were compared with an age-matched healthy control group (n=45). DCs were stained (Fascin) in circular sections of the internal thoracic artery taken during bypass surgery and mature (CD83^+^) and immature (CD209^+^) DCs were differentiated. Cell densities were obtained digitally. Descriptive statistics and correlation analysis generated hypotheses.

Circulating myeloid DCPs were reduced (p<0.001), concomitant CKD added no further reduction. Immature DCs were predominantly detected in the adventitia, mature DCs dominated in the intima (p<0.001). In CKD, reduced circulating myeloid DCPs correlated with immature DCs in the vessel wall (r=0.87; p=0.002) and rather mature DCs were detectable; in patients without CKD immature DCs were dominating. C-reactive protein correlated with mature DCs (r=0.38; p=0.026).

**Conclusions:** Quantities of DCPs and mature and immature DCs vary between the vessel wall layers. The dominance of immature myeloid DCs in the adventitia suggests activation of the vascular-associated lymphoid tissue (VALT). Intensified inflammation associated with CKD may have promoted the maturation of myeloic DCs.

## Introduction

As both antigen-presenting and antigen-processing cells, dendritic cells (DCs) represent an important junction between the innate and acquired immune system [1]. They are characterised by a high variability in function and morphology. DCs are the most effective antigen-presenting cells known and are of utmost importance involved in priming immune responses [2]. Most DCs evolve from a common CD34^+^ stem cell, except for localised follicular DCs [3]. Dendritic cell precursors (DCPs) are circulating in the bloodstream. Myeloid DCP (mDCP) and plasmacytoid DCP (pDCP) are differentiated according to their surface molecule pattern and their blood dendritic cell antigens (BDCA) [1, 4, 5]. DCPs are responsible for 1-2% of all circulating leucocytes in the peripheral blood [6]. They migrate through the blood stream to their destination targets, such as skin, lymphoid tissue or vessels, where they are able to differentiate into immature and mature DCs [7]. Whereas immature DCs are involved in antigen-detection and antigen-processing, mature DCs act as antigen-presenting cells and activate T-cells [8, 9]. The differentiation from immature to mature DCs is activated by inflammatory signals like tumour necrosis factor alpha (TNFα) and prostaglandin E2 [8].

Total counts of DCs were seen to be elevated in inflammatory diseases such as rheumatoid arthritis [8]. However, they can also be found increased in atherosclerotic plaques, attracted by several chemokines, cytokines and hypoxia following local inflammation [10]. Activated through damage-associated molecular patterns (DAMP) and pathogen-associated molecular patterns (PAMP), DCs co-stimulate further cells of the adaptive immune system, such as B-cells, T-cells, or natural killer cells [11, 12]. Its multifaceted role makes it an attractive immunomodulatory research target, including in the prevention and therapy of atherosclerosis [13].

Bobryshev et al. were the first to describe DCs in vascular walls [14]. Further studies provided evidence that DCs are especially increased in instable atherosclerotic plaques as compared to stable plaques [10]. Beyond that, DCPs circulating in the blood were shown to be reduced in patients with coronary artery disease (CAD) [15], possibly reflecting recruitment of DCPs in response to vascular damage and inflammation. Intima calcification was associated with atherosclerosis and inflammatory signals as macrophages were increasingly present [16]. Medial arterial calcification in CKD and diabetes mellitus is thought to reflect mineralisation rather than inflammation [17, 18].

Patients with chronic kidney disease (CKD) are at excess cardiovascular risk and thus suffer more frequently from acute coronary syndrome, left ventricular hypertrophy, and sudden cardiac death [19, 20]. In addition to common cardiovascular risk factors such as arterial hypertension, diabetes mellitus, dyslipidemia and adipositas, uremic factors like inflammation, oxidative stress, and overactivation of the sympathetic nervous system increase the cardiovascular risk of patients with CKD [21]. Although proinflammatory cytokines were increased in patients with CKD, DCPs were diminished in the peripheral blood [22, 23]. Consistent with that observation, in postmortem analysis of CKD patients, DCs were found to be increased in the aortic intima [24] which, in turn, may support the hypothesis that peripheral DCPs are decreased due to the recruitment of DCs in the vessel wall.

This study aimed to examine the relationship between reduced circulating DCP levels and DC counts in the vascular wall in patients with CAD undergoing bypass surgery. A particular focus was placed on patients with CKD compared with patients without CKD.

## Patients and Methods

### Patients

Between March 2016 and April 2017, a total of 63 patients with coronary artery disease (CAD) who underwent myocardial revascularisation using coronary artery bypass surgery at our university hospital were recruited. Patients with chronic inflammation or rheumatic diseases, malignancies, or with immunosuppression were excluded. All patients gave their written informed consent. The study was approved by the local ethics committee (4471-06/15). Baseline characteristics contained the following variables: age (years), sex (male/female), body-mass-index (kg/m²), arterial hypertension (mmHg), diabetes mellitus (%), cardiovascular familial predisposition (%), nicotine abuse (%), coronary artery disease (%), peripheral arterial occlusive disease (%), history of ischemic stroke (%), leukocytes (Gpt/L), thrombocytes (Gpt/L), C-reactive protein (mg/L), total cholesterine (mg/dL), creatinine (µmol/L), and estimated glomerular filtration rate (eGFR; mL/min/1.73 m²). The presence of CKD was defined as an eGFR ≤ 60ml/min/1.73m² at baseline.

A previously described local healthy cohort served as the comparison group after matching for age [23]. All patients in this control group had an eGFR > 60ml/min/1.73m² and CAD was excluded.

### Flow Cytometry

Immediately after admission to the hospital blood samples were collected and processed instantly after collection following the enumeration kit protocol (Miltenyi Biotec B.V. & Co. KG Bergisch Gladbach, Germany) [5], as previously described [21]. Thereby, samples were analysed using flow cytometry (FACS Calibur ^TM^; BD CellQuest ProTM software; Becton Dickinson GmbH, Germany, Heidelberg). The specific expression of BDCA markers for mDCP and pDCP enabled the determination of the total number of DCPs (tDCPs).

### Histological analyses of Arteria thoracica interna circular sections

During elective internal mammary artery bypass surgery, a circular section of the Arteria thoracica interna (Arteria mammaria) to be anastomosed was removed and preserved in 4% formaldehyde solution. Next, the samples were subjected to an automated tissue processor (Histokinette; Leica TP 1020, Leica Biosystem, Nussloch, Germany) and were paraffin embedded using a paraffin casting station (Leica EG1160, Leica Biosystem, Nussloch, Germany). Afterwards, 5µm sections were produced and stained with immunohistochemistry using the Catalysed Amplification System Kit and Catalysed Amplification System Kit II after experiencing delivery issues (AgilentDAKO, United States of America, Santa Clara, CA). The specific antibodies for mDCPs and pDCPs and dilutions used are given in **Table S1**. For visualisation of the cell nuclei the sections were additionally stained with hematoxylin for 2 minutes.

The target cells were quantified digitally using ImageJ software (National Institutes of Health, Bethesda, MD, USA) at 100x magnification. To minimise errors caused by local variations in cell populations, the images were analysed separately for each quadrant of the circumference, followed by averaging.

### Statistical Analyses

Statistical analyses were performed using Microsoft Excel 2010 (Microsoft Corporation, Redmond, WA, USA) and IBM SPSS Statistics (version 30.0.0.0 (171)) (IBM Corporation, Armonk, NY, USA). Differences between two groups were analysed using Mann-Whitney-U test and chi-square tests, as appropriate. Correlations between non-normally distributed variables were assessed using Spearman’s rank correlation coefficient (Spearman’s-Rho). P-value of *P* < 0.05 was considered statistically significant.

## Results

### Study population

A total of n=63 patients were primarily included, of whom 59 patients had valid DCP counts that could be determined and compared with 45 age-matched controls. Tissue samples of the arteria thoracica interna for histological staining were obtained from only 38 patients due to technical reasons (e.g. change of surgical technique, limited vessel diameter, surgeon’s decision). Following preparation for immunohistochemical staining 35 samples (34 for CD209 and CD38) were available for the final analyses (**Figure S1**).

Baseline characteristics of all 59 patients with CAD (study cohort) and the age-matched control group of 45 patients are displayed in **Table 1**. The study cohort contains more frequently patients with familial predisposition for CAD, nicotine abuse, and CAD-associated comorbidities such as peripheral artery occlusive disease, diabetes mellitus, and impaired kidney function. Within the whole study cohort, 20 patients (34%) had an eGFR ≤ 60mL/min/1.73m² and were classified as the CKD subgroup. The mean eGFR in this CKD subgroup was 35.6±19.4 mL/min/1.73m², compared with 78.6±12.3 mL/min/1.73 m² in patients without CKD (p < 0.001; **Table S2**).

**Table 1.** Baseline characteristics of the study group and control group.

|  | *Study cohort*<br>*with CAD\**<br>*n=59* | *Study cohort with CAD*<br>*and tissue sample*<br>*n=38* | *Control group\**<br>*n=45* | *p-value\** |
| --- | --- | --- | --- | --- |
| *Age [years]* | 68.5 ± 8.4 | 68.2 ± 8.2 | 65.7 ± 7.1 | 0.113 |
| *Female [%]* | 12 (20.3) | 12 (20.3) | 27 (60.0) | **<0.001** |
| *Body-mass-index [kg/m<sup>2</sup>]* | 29.4 ± 4.9 | 30 ± 4.9 | 27.8 ± 4.6 | 0.131 |
| *Adipositas (BMI>30 kg/m<sup>2</sup>) [%]* | 25 (42.4) | 17 (44.7) | 23 (51.1) | 0.376 |
| *Arterial hypertension [%]* | 56 (94.9) | 36 (94.7) | 39 (86.7) | 0.138 |
| *Diabetes mellitus [%]* | 25 (42.4) | 16 (42.1) | 7 (15.6) | **0.003** |
| *Familial predisposition [%]* | 41 (69.5) | 29 (76.3) | 12 (26.7) | **<0.001** |
| *Nicotine abuse [%]* | 34 (57.6) | 24 (63.2) | 11 (24.4) | **<0.001** |
| *Coronary artery disease [%]* | 59 (100) | 38 (100) | 0 (0) | **<0.001** |
| *Peripheral arterial occlusive*<br>*disease [%]* | 10 (16.9) | 6 (15.8) | 0 (0) | **0.004** |
| *History of ischemic stroke [%]* | 9 (15.3) | 4 (10.5) | 3 (6.7) | 0.174 |
| *Leukocytes [Gpt/l]* | 7.4 ± 1.7 | 9.5 ± 11.3 | 7.1 ± 1.4 | 0.231 |
| *Thrombocytes [Gpt/l]* | 237 ± 78 | 220 ± 90 | 227 ± 46 | 0.201 |
| *C-reactive protein [mg/l]* | 7.0 ± 21.5 | 6.3 ± 19.7 | 2.6 ± 3.6 | 0.823 |
| *Total cholesterine [mg/dl]* | 4.5 ± 1.1 | 4.5 ± 1.2 | 5.8 ± 1.3 | **<0.001** |
| *Creatinine [μmol/l]* | 134 ± 178 | 153 ± 219 | 78 ± 17 | **<0.001** |
| *Estimated glomerular filtration*<br>*rate [ml/min/1.73m<sup>2</sup>]* | 65.3 ± 25.0 | 65.0 ± 25.0 | 79.0 ± 15.2 | **0.006** |

Numbers in mean ± standard deviation or n (%).
CAD: coronary artery disease; BMI: body-mass-index.

### Dendritic cell precursors in the peripheral blood

In FACS analysis, both the absolute total DCP count and the absolute myeloid DCP count were significantly diminished in patients with CAD (n=59) as compared to controls (p < 0.001, **Figure 1, Table S3**). No difference in densities of circulating DCPs was monitored between patients with and without CKD (**Table S3**).

**Figure 1.**
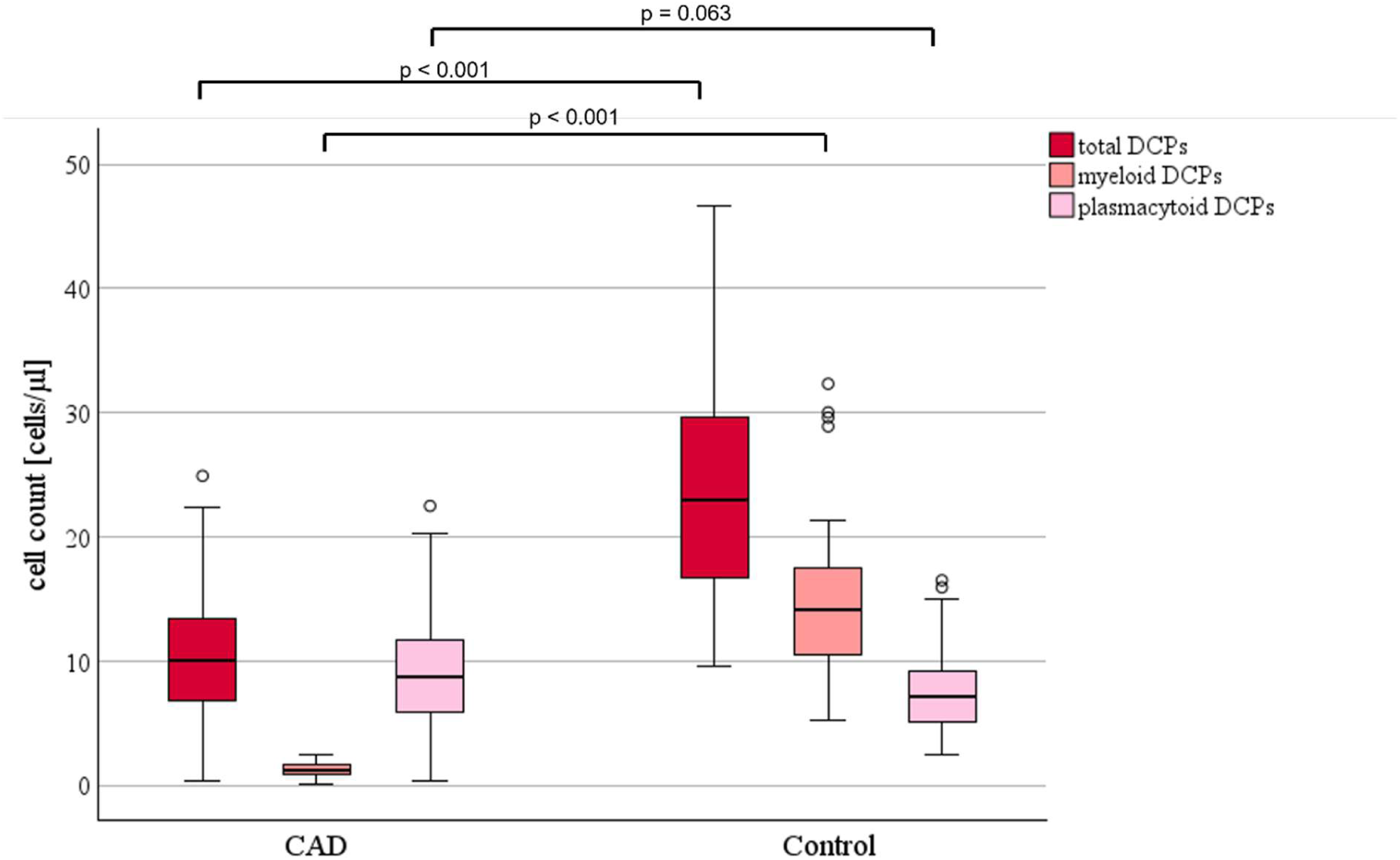
Cell counts of dendritic cell precursors (DCP; total, myeloid, plasmacytoid) between the study group with coronary artery disease (CAD; n=59) and the control group (n=45)

Of note, tDCP (9.6 ± 6.8 / µl) and mDCP (1.2 ± 5.0 / µl) levels were significantly reduced in patients with diabetes mellitus compared to patients without diabetes mellitus (15.9 ± 9.7 and 6.3 ± 8.6 / µl; p = 0.001 and p = 0.002).

Within the patients with a tissue sample of the internal thoracic artery (n=38) the results were consistent as tDCP and mDCP counts were significantly reduced compared to the control group (**Table 2**).

**Table 2.** Dendritic cell precursors in the peripheral blood in patients with CAD in comparison to the control group with successful collection of a tissue sample.

|  | *Study cohort with CAD*<br>*and tissue sample*<br>*n=38* | *Control group*<br>*n=45* | *p-value* |
| --- | --- | --- | --- |
| *tDCP absolute (per μL)* | 9.65 ± 5.06 | 23.52 ± 8.65 | **< 0.001** |
| *tDCP relative (%WBC)* | 0.26 ± 0.12 | 0.33 ± 0.10 | **0.009** |
| *mDCP absolute (per μL)* | 1.15 ± 0.56 | 14.64 ± 6.69 | **< 0.001** |
| *mDCP relative (% WBC)* | 0.17 ± 0.09 | 0.20 ± 0.07 | **0.035** |
| *pDCP absolute (per μL)* | 8.50 ± 4.56 | 7.97 ± 3.66 | 0.579 |
| *pDCP relative (%WBC)* | 0.09 ± 0.05 | 0.12 ± 0.06 | 0.071 |

Numbers in median (Interquartile range), mean ± standard deviation or n (%).
CAD: coronary artery disease; t/m/pDCP: total/myeloid/plasmacytoid dendritic cell precursors; abs/rel: absolute/relative count; WBC: white blood count

### Dendritic cells (DC) in the vessel wall

The tissue samples of the patients with CAD were stained for fascin (myeloid DC), CD83 (mature myeloid DC) and CD209 (immature myeloid DC). Representative images are shown in **Figure 2**.

**Figure 2.**
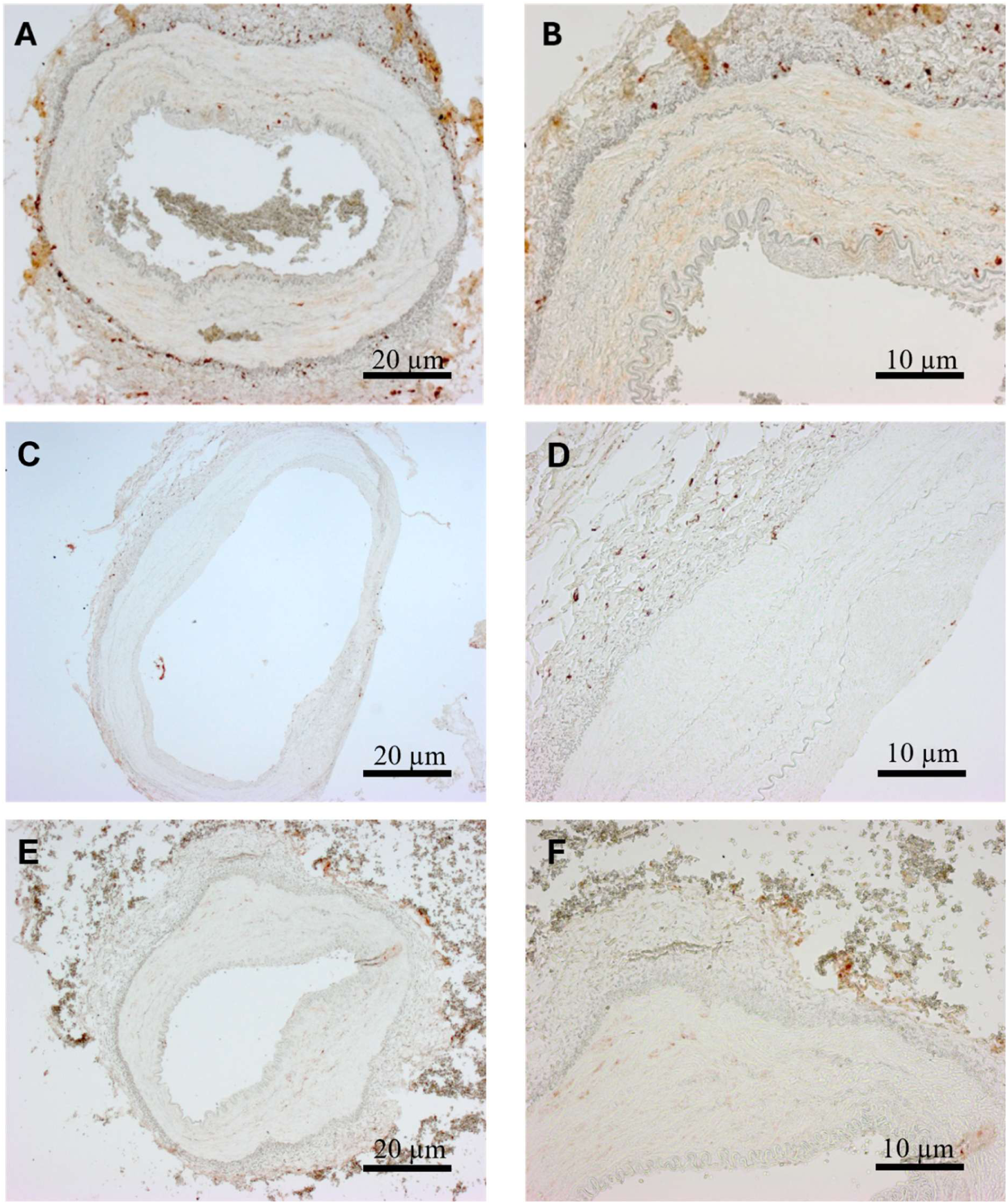
Immunohistochemical detection of Fascin (A, B), CD209 (C, D), and CD83 (E, F) in the internal thoracic artery. Panels A, C, and E are shown in 50x magnification, whereas panels B, D, and F are shown at 100x magnification.

The distribution of DCs across the different layers of the vessel wall is displayed in **Figure 3**, and their quantities are summarised in **Table S4**. Overall, staining showed the highest DCP-density in the adventitia, followed by the intima (p < 0.001). While immature CD209^+^-DCs predominated in the adventitia, mature CD83^+^-DCs were more abundant in the intima. Patients with CKD tended to have lower fascin-positive and CD209^+^-DC densities compared to patients without CKD, but a higher CD83-positive DC density (**Figure 3, Table S4**).

**Figure 3.**
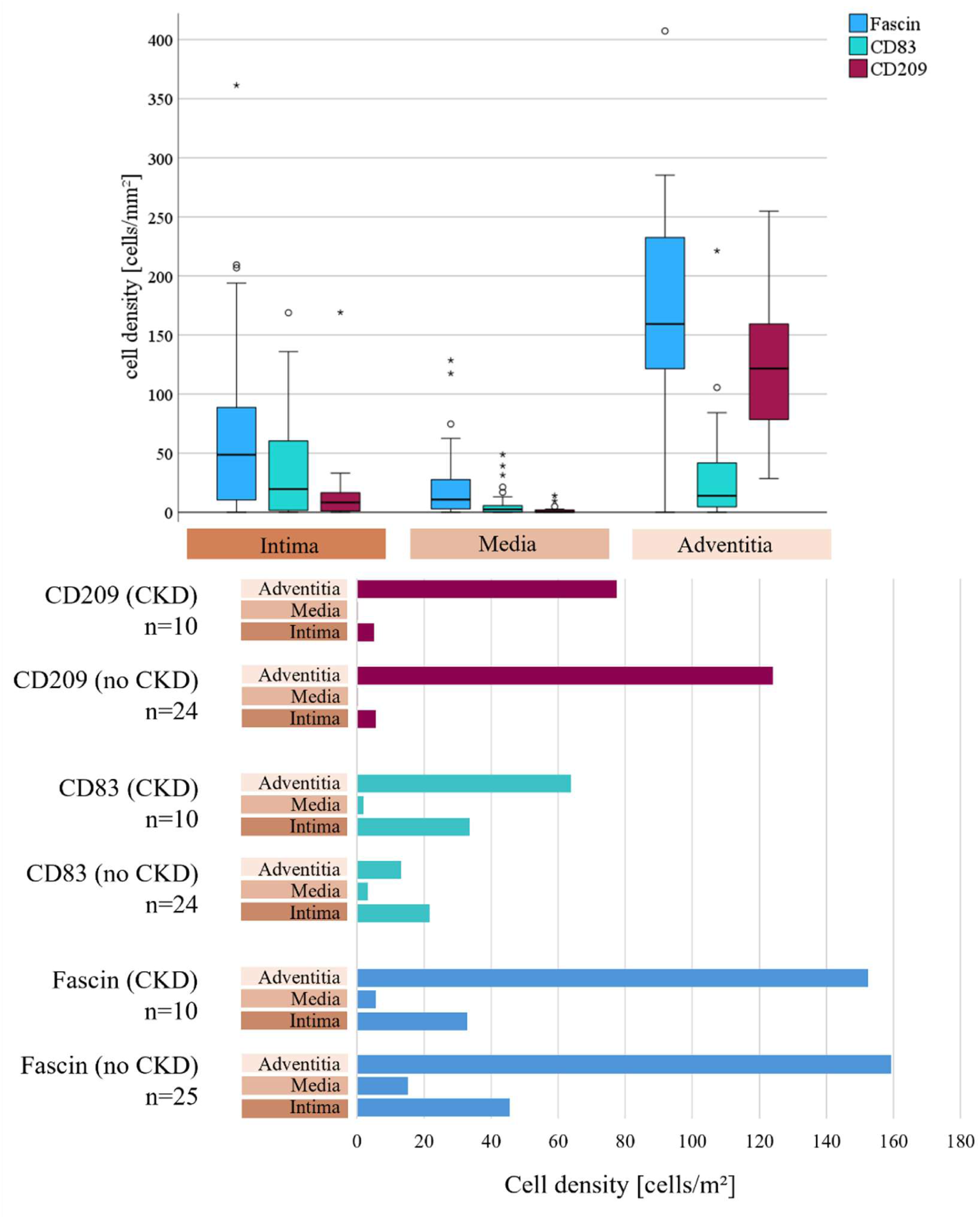
Dendritic cell (DC) densities in the vessel wall layers of the internal thoracic artery of n=35 patients with coronary artery disease (boxplot diagram; Kruskal-Wallis p < 0.001 for Fascin (all DCs), CD83 (mature DCs), and CD209 (immature DCs)) and juxtaposition of DC densities in the subgroups of patients with and without chronic kidney disease (CKD).

Fascin- and CD83-positive cell densities in the intima were positively correlated to C-reactive protein (r_Fascin_ = 0.352, p = 0.038; r_CD83_ = 0.381, p = 0.026). Correlation analyses were not significant for leucocyte counts in the blood or the presence of diabetes mellitus.

Stainings for plasmacytoid DCs (CD309 and CD123) did not yield meaningful results, as they resulted in either ubiquitous staining or merely nonspecific staining.

### Relationship between DCPs of the peripheral blood and DCs of the vessel wall

In the total study cohort with successful staining (n=38), no direct correlation between DCPs in the peripheral blood and DC density in the layers of the internal thoracic artery was found. However, in the subgroup of patients with CKD, immature mDCs (CD209^+^) in the intima were significantly positively associated with the relative number of circulating tDCPs (*r*_s_ =0.870, *p*=0.002), mDCPs (*r*_s_= 0.868, *p*=0.002), and pDCPs (*r_s_*=0.780, *p*=0.013). By analogy, in patients without CKD, a negative correlation was found for immature mDCs (CD209^+^) in the intima for counts of circulating tDCPs (*r_s_*= -0.590, *p*=0.005), mDCPs (*r_s_*= -0.610, *p*=0.003), and pDCPs (*r_s_*= -0.571, *p*=0.007).

Similarly, fascin-positive cells could be positively correlated with relative counts of tDCPs (*r_s_*=0.733, *p*=0.025) and pDCPs (r= 0.793, *p*=0.011) in the average of all layers of the vessel wall, and in particular in the intima (tDCP *r_s_*= 0.686, *p*=0.041), in patients with CKD.

## Discussion

Inflammation is an important commonality in patients with cardiovascular diseases and chronic kidney disease (CKD) and is a well-known independent mortality risk factor [25]. Inflammatory processes, including the activation of both the innate and acquired immune system, are involved in the development of atherosclerosis [26]. This study aims to better understand immunological processes that may be associated with vascular inflammation in high-risk patients with confirmed cardiovascular disease with and without chronic kidney disease. Typical risk factors were observed more frequently within the study cohort of 59 patients undergoing coronary bypass surgery, such as smoking and a positive family history for coronary artery disease (CAD), alongside related comorbidities such as peripheral artery occlusive disease, diabetes mellitus and chronic kidney disease [27, 28]. The prevalence of 34% of patients with CKD in our study cohort can be considered representative, as cross-sectional data from 27 European countries describe a prevalence of CKD in patients with CAD of 27.3% [29].

The absolute count of total dendritic cell precursors (tDCP) in the blood and, in particular, the absolute count of myeloid DCP (mDCP) was reduced in the study cohort of patients with severe CAD compared with the coronary healthy and age-matched controls. Notably, the count of plasmacytoid DCPs (pDCPs) was unchanged. The affection of dendritic cells (DC) and their precursors by vascular diseases was shown previously [15, 26]. In patients with angina pectoris and acute myocardial infarction mDCPs in the blood were reduced, but not pDCPs [15, 30]. By this, our findings on the circulating DC precursors are mainly in line with previous work in this field; however, in a larger study cohort also pDCPs were reduced in a subgroup of patients with severe CAD, assessed by a scoring system regarding the maximum grade of coronary stenosis in each of the 15 coronary segments [31].

The severity of chronic kidney disease (CKD) is additionally known to reduce mDCPs and pDCPs in the circulating blood [23, 32, 33], with the reduction in DCPs being more pronounced in patients with a positive history of cardiovascular events [21]. In the present study, with – for methodological reasons – only a limited number of participants included, the counts of tDCPs tended to be somewhat lower in patients with CAD and CKD compared to patients with CAD without CKD, however, statistical significance was not achieved. Since all patients suffered from severe CAD and as the levels of DCPs in these patients were already reduced, the additional effect due to CKD might have been too small for the sample size of twenty patients (regarding precursors) or ten patients (regarding DCs in the vessel wall).

Moreover, hyperglycaemia can induce chronic vascular inflammation mediated by various pathways including those mediated by DCs [34]. Patients with diabetes mellitus in our study cohort had significantly lower levels of tDCPs and mDCPs compared to patients without diabetes mellitus confirming previous findings [23].

The reduced quantity of DCPs is assumed to be mainly driven by vascular migration of DCPs in surrounding tissues, including the vessel wall, following specific chemokines [7, 30, 31]. We hypothesised that immature (and mature) DCs would be detectable more frequently in the vessel wall, correlating to inflammation and reduced quantity in the peripheral blood as a consequence of migration. In uremic conditions, DCPs are further impaired in their functionality [35]. A reversed migration from the vessel wall towards the bloodstream is suppressed by platelet-activating factor (PAF) and low-density lipoprotein (LDL) [36]. Our goal was to determine, whether in high-risk patients with CAD, reduced concentrations of circulating DCPs are associated with the number of DCs that have migrated into the vessel wall. A direct comparison of mDCs in the vessel wall between patients with CAD and healthy control subjects was not possible due to the understandable absence of tissue samples from healthy individuals.

To detect migrated myeloid DCs in the vessel wall, circular tissue samples of the internal thoracic artery of patients with CAD were stained for Fascin (myeloid DC), CD83 (mature DC), and CD209 (immature DC). In patients without vascular diseases (and without CKD), migrated DCs were previously found to be mainly in the intima following microinflammation and mechanical shear forces [37, 38]. In addition, in close relation to local vasa vasorum, DCs were also detected in the adventitia [38]. In this present study, immunohistochemical staining and quantification of DCs in patients with CAD revealed a predominant distribution of fascin- and CD209^+^-DCs in the adventitia and, to a lesser extent, in the intima. Hematopoietic-derived DCs in the adventitia were considered as part of the vascular-associated lymphoid tissue (VALT) when regarding aortic aneurysms [3, 39]. The primary objective of the VALT is to screen the perivascular tissue for potential harmful antigens and is mainly located in the arterial intima, especially in arteriosclerotic plaque regions and in the adventitia close to vasa vasorum [40, 41]. In this regard the VALT is part of tertiary lymphoid structures (TLS). The TLS is defined as lymphoid aggregates that develop in non-hematopoietic organs during cancer or chronic inflammation of various causes [3, 42]. A major focus within the investigation of the TLS is on the development of therapeutic targets in autoimmune diseases. However, also CAD and CKD reflect a state of chronic inflammation. In fact, the cellular dimension of the TLS is known to correlate strongly with lesion size, plaque instability, and rupture in patients with CAD [43]. Furthermore, TLS may suppress regeneration of kidney tissues in the context of age-related progression of CKD [42]. This study hints to the fact that chronic vascular inflammation in the context of CAD may activate the VALT.

Mature, CD83^+^-DCs were found more pronounced in the intima as compared to immature CD209^+^-DCs and were equally distributed between the intima and adventitia. CD83^+^ cells are known for their activation of T-cells and leucocytes and can serve as a marker for vascular inflammation [44]. Taking this together it is plausible that mature DCs are found in the intima as inflammation leads to migration of DCs from the vessel’s lumen; however, additional migration may take place via the local vasa vasorum.

Patients with CAD *and* CKD tended to have a higher mature DC count (CD83^+^) in both intima and adventitia compared to patients with CAD but without CKD. It seems conceivable that statistical tests were unable to demonstrate statistical significance in these subgroups due to the limited sample size.

In addition, patients with CAD and CKD tended to lower absolute DC densities (Fascin-positive) and furthermore showed a co-directional (positive) correlation of reduced DCP and diminished immature (CD209^+^) DC counts in the intima. Possibly these observations can be related to uremic effects, which is known to lead to a depletion and downregulation of dendritic cells [23, 32, 45]. On the other hand, in patients with CAD but *without* CKD, solely vascular inflammation may be dominating, as we observed reduced DCPs but higher counts of immature DCs in the vessel wall with an inverse (negative) correlation for the intima cell density.

Moreover, Fascin- and CD83^+^-DC counts in the intima were positively correlated to the nonspecific inflammation marker C-reactive protein (CRP). Patients with CAD and CKD tended to have higher CRP values. Furthermore, CRP is known to induce DC activation and maturation [30]. These considerations lead to the thesis that chronic vascular inflammation, intensified by CKD, promotes maturation of immature DCs (CD209^+^) to mature DCs (CD83^+^) in the vessel wall. Whereas immature DCs have a reduced potential to release pro-inflammatory cytokines, mature DCs have diminished potential to express MHC- and costimulatory molecules for antigen presentation [46]. Previous findings revealed that patients without chronic inflammation have a reduced mature, CD83-positive DC count in the intima which underscores our assumptions [47, 48]. In patients with vascular inflammation such as CAD, however, both CD83^+^-DCs and Fascin-positive DCs were increased, especially in arteriosclerotic plaque regions [10, 49]. Usually, the internal thoracic artery is not prone to arteriosclerotic plaque formation, which is beneficial for bypass surgery, however, this may have diminished the significance of our study results in this regard [50]. As a consequence, active remodelling, as described in the formation of arteriosclerotic plaques, cannot be observed based on the material examined [49, 51].

This present study is, to our best knowledge, the first evaluation of plaque-free arterial tissue regarding DCs in patients with CAD with and without CKD analysing tissue samples taken from living subjects. However, it is limited mainly in the size of the study cohort, where both tissue collection and staining were successful as well as by the localization of the tissue sample taken. Furthermore, the association between DCPs in the blood and DCs in the vessel wall could only be investigated within the group of patients with CAD as from a healthy control group for ethical reasons no vascular tissue samples could be obtained.

In summary, our findings confirm that in patients with CAD circulating DCPs are diminished. Immature DCs in the vascular wall were predominantly located in the adventitia, whereas mature DCs were predominant in the intima. A direct relation as a consequence of migration between lower DCPs in the peripheral blood and higher counts of DCs in the vessel wall could not be monitored, but the increased cell density of DCs in the adventitia suggests an activation of the VALT as a consequence to chronic vascular inflammation. CKD may promote maturation of DCs in the intima which appears to be linked to the state of intensified chronic inflammation.

## Data Availability

The data that support the findings of this study are available from the corresponding author, JR, upon reasonable request

## Author contributions

JR wrote the manuscript. SB and JR performed statistical analysis. Conceptualisation was outlined by MB and SB. SB coordinated and collected the blood and tissue samples for FACS-analysis, staining, and cell counts. TB and EO ensured correct laboratory methodology, GF and TD provided and conducted the tissue sample collection. DK, IR and GW were supervising and provided expert knowledge in the field. SB, TB, TD, GW, and MB revised the manuscript. All authors read and approved the final version of the manuscript.

## Acknowledgments

We would like to extend our special thanks to the Cardiothoracic Surgery team for their cooperation and for providing the tissue samples. We would like to thank for the excellent technical assistants to Elke Oswald (Department of Internal Medicine III, Nephrology) and Katja Schubert (Department of Internal Medicine I, Cardiology and Angiology).

## Supplement

**Figure S1.**
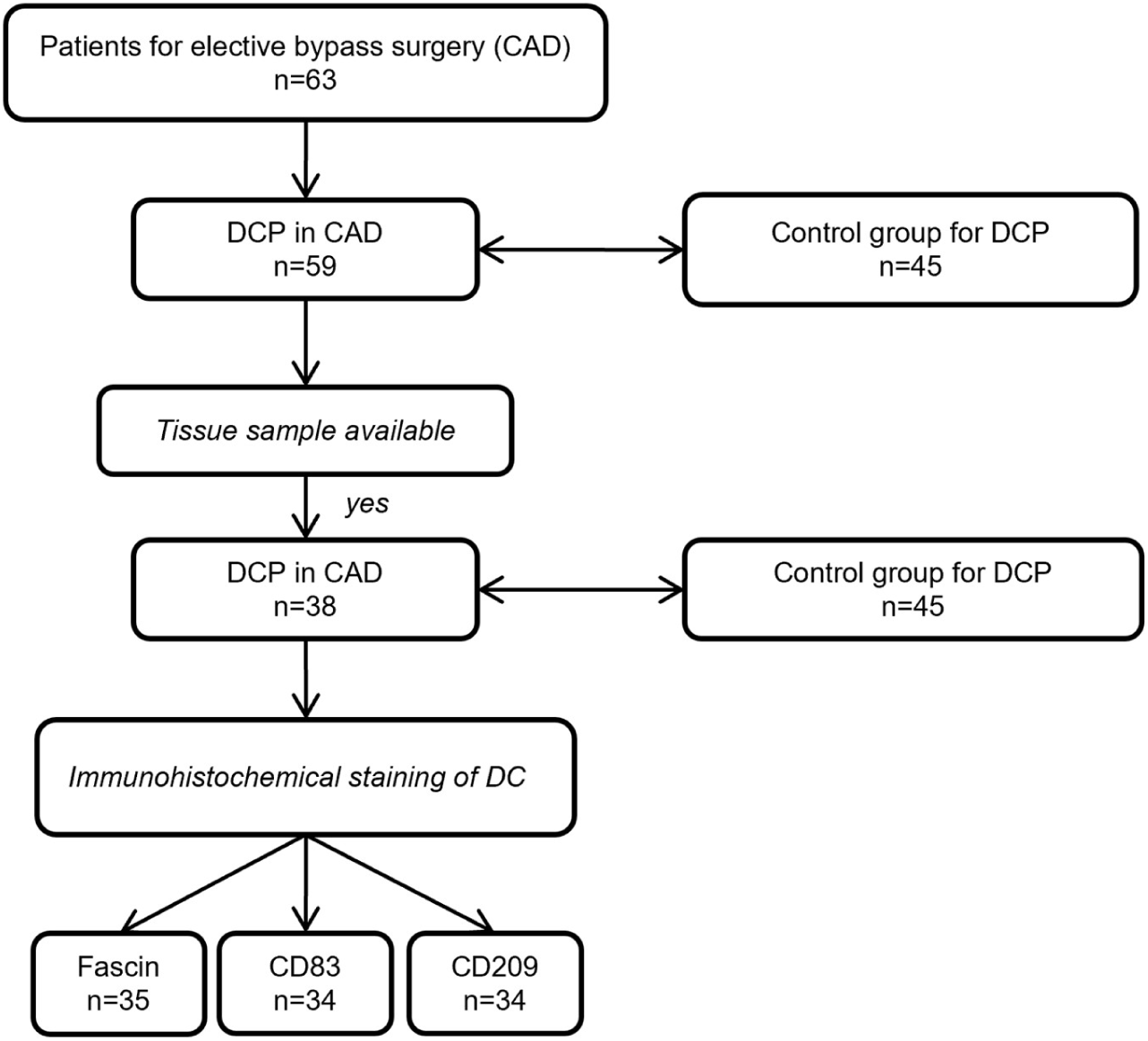
Enrolment flow chart; CAD: coronary artery disease; DCP: dendritic cell precursors; DC: dendritic cells.

## Tables

**Table S1.**
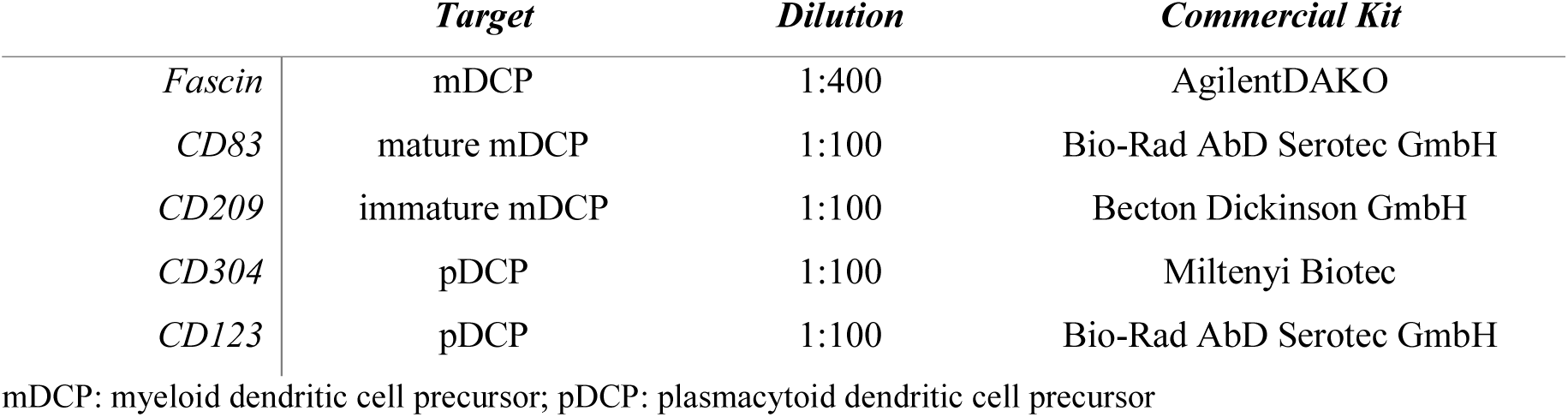
Commercial Kits used for the analysed target cells.

**Table S2.** Comparison of baseline characteristics between patients with CAD with and without CKD with successful collection of a tissue sample.

|  | *CAD with CKD*<br>*n=12* | *CAD without CKD*<br>*n=26* | *p-value* |
| --- | --- | --- | --- |
| *Age [years]* | 68.7 ± 9.7 | 68.0 ± 7.6 | 0.860 |
| *Female [%]* | 5 (25.0) | 7 (17.9) | 0.850 |
| *Body-mass-index [kg/m<sup>2</sup>]* | 32.7 ± 5.9 | 28.6 ± 3.8 | 0.052 |
| *Adipositas (BMI>30kg/m<sup>2</sup>) [%]* | 7 (58.3) | 10 (38.5) | 0.252 |
| *Arterial hypertension [%]* | 12 (100.0) | 24 (92.3) | 0.324 |
| *Diabetes mellitus [%]* | 8 (66.7) | 8 (30.8) | **0.037** |
| *Familial predisposition [%]* | 8 (66.7) | 21 (80.8) | 0.342 |
| *Nicotine abuse [%]* | 8 (66.7) | 16 (61.5) | 0.761 |
| *Statine therapy [%]* | 11 (91.7) | 22 (84.6) | 0.550 |
| *Coronary artery disease [%]* | 12 (100.0) | 26 (100.0) | >0.999 |
| *Peripheral arterial occlusive disease [%]* | 3 (25.0) | 3 (11.5) | 0.290 |
| *History of ischemic stroke [%]* | 2 (16.7) | 2 (7.7) | 0.402 |
| *Leukocytes [Gpt/l]* | 8.3 ± 2.2 | 10.1 ± 13.6 | 0.293 |
| *Thrombocytes [Gpt/l]* | 225.0 ± 90.7 | 216.9 ± 91.1 | 0.660 |
| *C-reactive protein [mg/l]* | 13.7 ± 32.1 | 5.4 ± 17.5 | 0.215 |
| *Total cholesterine [mg/dl]* | 4.3 ± 0.9 | 4.7 ± 1.3 | 0.638 |
| *Creatinine [μmol/l]* | 305.4 ± 351.6 | 82.9 ± 13.5 | **<0.001** |
| *Estimated glomerular filtration rate*<br>*[ml/min/1.73m<sup>2</sup>]* | 35.6 ± 19.4 | 78.6 ± 12.3 | **<0.001** |

Numbers in mean ± standard deviation or n (%).
CAD: coronary artery disease; CKD: chronic kidney disease; BMI: body-mass-index.

**Table S3.** Dendritic precursors in the peripheral blood in patients with CAD in comparison to the control group and with and without CKD.

|  | *CAD*<br>*n=59* | *Control group*<br>*n=45* | *p-value* | *CAD and CKD*<br>*n=20* | *CAD without CKD*<br>*n=39* | *p-value* |
| --- | --- | --- | --- | --- | --- | --- |
| *tDCP abs.*<br>*(per μl)* | 10.02 (0.39/24.91)<br>10.92 ± 5.26 | 23.00 (9.60/46.61)<br>23.52 ± 8.65 | < **0.001** | 9.62 (0.39/22.39)<br>10.57 ± 4.93 | 10.35 (2.29/24.91)<br>11.10 ± 5.47 | 0.962 |
| *tDCP rel.*<br>*(% WBC)* | 0.29 (0.01/0.53)<br>0.29 ± 0.12 | 0.33 (0.15/0.59)<br>0.33 ± 0.10 | 0.069 | 0.29 (0.01/0.53)<br>0.28 ± 0.13 | 0.27 (0.09/0.51)<br>0.29 ± 0.11 | 0.873 |
| *mDCP abs.*<br>*(per μl)* | 1.19 (0.06/2.41)<br>1.30 ± 0.57 | 14.11 (5.22/32.32)<br>14.64 ± 6.69 | < **0.001** | 1.16 (0.06/2.11)<br>1.23 ± 0.53 | 1.22 (0.45/2.42)<br>1.34 ± 0.59 | 0.608 |
| *mDCP rel.*<br>*(% WBC)* | 0.18 (0.01/0.37)<br>0.19 ± 0.08 | 0.20 (0.08/0.38)<br>0.20 ± 0.07 | 0.228 | 0.18 (0.01/0.37)<br>0.17 ± 0.09 | 0.18 (0.05/0.36)<br>0.19 ± 0.08 | 0.586 |
| *pDCP abs.*<br>*(per μl)* | 8.67 (0.34/22.49)<br>9.61 ± 4.74 | 7.20 (2.45/16.50)<br>7.97 ± 3.66 | 0.063 | 8.50 (0.34/20.76)<br>9.34 ± 4.49 | 8.71 (1.73/22.49)<br>9.76 ± 4.91 | 0.923 |
| *pDCP rel.*<br>*(% WBC)* | 0.09 (0.00/0.25)<br>0.10 ± 0.051 | 0.11 (0.04/0.30)<br>0.12 ± 0.06 | 0.308 | 0.10 (0.0/0.20)<br>0.11 ± 0.06 | 0.09 (0.04/0.25)<br>0.10 ± 0.05 | 0.664 |

Numbers in median (Interquartile range), mean ± standard deviation or n (%).
CAD: coronary artery disease; CKD: chronic kidney disease; t/m/pDCP: total/myeloid/plasmacytoid dendritic cell precursors; abs/rel: absolute/relative count; WBC: white blood count

**Table S4.** Distribution of dendritic cell densities in patients with CAD with and without CKD.

|  | *Total*<br>*n=35\** | *CAD with CKD\**<br>*n=10* | *CAD without CKD\**<br>*n=25* | *p-value\** |
| --- | --- | --- | --- | --- |
| *Fascin Intima*<br>(cells/mm<sup>2</sup>) | 45.6 (80.2)<br>66.8 ± 79.5 | 32.9 (114.5)<br>80.8 ± 117.3 | 45.6 (75.1)<br>61.2 ± 60.6 | 0.798 |
| *Fascin Media*<br>(cells /mm<sup>2</sup>) | 11.1 (25.8)<br>22.7 ± 30.8) | 5.6 (21.8)<br>21.0 ± 39.0 | 15.3 (25.3)<br>23.4 ± 27.8 | 0.371 |
| *Fascin Adventitia*<br>(cells /mm<sup>2</sup>) | 159.3 (116.1)<br>181.7 ± 128.3 | 152.4 (105.5)<br>133.7 ± 72.0 | 159.3 (115.6)<br>200.8 ± 141.4 | 0.235 |
| *Fascin Total*<br>(cells /mm<sup>2</sup>) | 80.0 (81.6)<br>90.4 ± 59.3 | 70.9 (81.4)<br>78.5 ± 59.4 | 80.0 (82.4)<br>95.1 ± 59.8 | 0.454 |
| *CD83 Intima*<br>(cells/mm<sup>2</sup>) | 26.0 (67.5)<br>60.0 ± 132.4 | 33.6 (107.8)<br>117.4 ± 232.8 | 21.6 (63.63)<br>36.1 ± 43.5 | 0.341 |
| *CD83 Media*<br>(cells/mm<sup>2</sup>) | 3.03 (8.6)<br>7.7 ± 11.8 | 2.0 (17.8)<br>8.3 (11.1) | 3.3 (5.7)<br>7.5 ± 12.3 | 0.809 |
| *CD83 Adventitia*<br>(cells/mm<sup>2</sup>) | 18.0 (54.3)<br>34.3 ± 44.1 | 63.8 (75.8)<br>49.1 ± 38.6 | 13.2 (29.2)<br>28.1 ± 45.6 | 0.159 |
| *CD83 Total*<br>(cells/mm<sup>2</sup>) | 21.5 (32.5)<br>34.0 ± 52.4) | 41.5 (63.6)<br>58.2 ± 83.7 | 16.0 (24.3)<br>23.1 ± 29.0 | 0.223 |
| *CD209 Intima*<br>(cells/mm<sup>2</sup>) | 5.7 (15.2)<br>13.7 ± 29.2 | 5.1 (13.4)<br>8.2 ± 10.4 | 5.7 (17.7)<br>16.0 ± 34.1 | 0.513 |
| *CD209 Media*<br>(cells/mm<sup>2</sup>) | 0.3 (1.8)<br>1.4 ± 2.9 | 0.2 (2.4)<br>1.7 ± 3.0 | 0.3 (1.6)<br>1.3 ± 2.9 | 0.733 |
| *CD209 Adventitia*<br>(cells/mm<sup>2</sup>) | 117.3 (86.4)<br>118.4 ± 58.0 | 77.5 (106.8)<br>106.0 ± 61.4 | 124.0 (76.0)<br>123.6 ± 57.0 | 0.326 |
| *CD209 Total*<br>(cells/mm<sup>2</sup>) | 30.7 (24.3)<br>33.4 ± 19.4 | 20.6 (27.6)<br>29.0 ± 16.7 | 33.6 (20.9)<br>35.2 ± 20.4 | 0.290 |

Numbers in median (Interquartile range), mean ± standard deviation or n (%).
CAD: coronary artery disease; CKD: chronic kidney disease; BMI: body-mass-index.

